# National burden and optimal vaccine policy for Japanese encephalitis virus in Bangladesh

**DOI:** 10.1101/2025.07.07.25330995

**Authors:** Mariana Perez Duque, Kishor K. Paul, Rebeca Sultana, Gabriel Ribeiro dos Santos, Megan O’Driscoll, Abu M. Naser, Mahmudur Rahman, Mohammad Shafiul Alam, Hasan M. Al-Amin, Mohammed Z. Rahman, Mohammad E. Hossain, Repon C. Paul, Elias Krainski, Stephen P. Luby, Simon Cauchemez, Jessica Vanhomwegen, Emily S. Gurley, Henrik Salje

## Abstract

**Background:** Bangladesh first reported Japanese encephalitis virus (JEV) in 1977 and has seen regular cases since, however, no JEV vaccination program currently exists. A barrier to the use of JEV vaccines has been a limited understanding of the underlying burden.

**Methods:** We conducted a nationally representative serological community study in 70 communities in individuals of all ages (N=2,938, October 2015-January 2016). Serum samples were tested for IgG antibodies against JEV. We developed spatially explicit serocatalytic models to estimate the underlying force of infection across the country. We then used mathematical models to estimate the annual JE disease burden currently and under different vaccination strategies.

**Findings:** The overall JEV seroprevalence in Bangladesh was 3.4% (95%CI: 2.8–4.1, range 0–28% across communities). The annual probability of infection was 0.005 (95%CI: 0.003–0.009), with risk greatest near border regions. We estimated that annually there are 157 clinical cases (95%CI: 89-253) and 31 deaths (95%CI: 18-52). A vaccination strategy in the 10 most affected districts in 60% of 1–15 year olds would require 5 million doses and avert 1 case per 100,000 doses over five years compared to 35 million doses and 0.5 cases averted for a nationwide campaign. No vaccination scenario was cost-effective under a willingness-to-pay of three-times gross domestic product.

**Interpretation:** A spatially targeted vaccine campaign would be most effective in reducing JEV burden, however, would still not meet standard cost effectiveness targets.

**Funding:** CDC

## Introduction

Japanese encephalitis virus (JEV) is endemic across South and Southeast Asia and is the most common cause of viral encephalitis in Asia^1^. The virus circulates in a zoonotic cycle mostly between mosquitoes, wading birds and pigs^2^. Humans become infected by but cannot infect mosquitoes^3^. In consequence ecological control of JEV is unrealistic and human vaccination is the main public health tool to prevent the disease. Bangladesh first reported JEV in 1977 and has seen regular cases since the start of hospital surveillance in 2003.^4,5^ Bangladesh has obtained provisional Gavi support to introduce JE vaccination. However, the optimal strategy to roll out the vaccine and the associated health impacts are unclear.

Most JEV infections are asymptomatic or cause mild disease. However, between one in 250 and one in 1000 develop symptomatic Japanese encephalitis (JE), usually an acute meningoencephalitis syndrome with high fever, headache, neck stiffness, seizures, paralysis, similar to other neurotropic flaviviruses, such as West Nile fever.^3,6^ The case-fatality ratio among hospitalised patients is estimated to be 20-30%, and long-term sequelae, such as cognitive impairment and motor deficits occur in 30-50% among those who survive^7^. The economic burden of a JE acute hospitalised episode is estimated to be high and often paid out of pocket by patients and their families, which can come from savings and incur in sustained debt.^8^

JEV vaccines have been available for decades and are highly effective against the disease with long-lasting immunity.^9^ The World Health Organization (WHO) recommends vaccination in endemic countries where JE constitutes a public health priority. Since humans are dead-end hosts, human vaccination has no impact on the zoonotic transmission cycle. Therefore, even when JE cases are low, vaccination should be considered in areas that provide a suitable environment for viral transmission. A critical barrier in the use of JEV vaccines in Bangladesh has been a limited understanding of where JEV circulates. This knowledge gap has been driven by the high proportion of clinically inapparent infections, frequent misdiagnosis, the difficulty in confirmatory testing that requires cerebrospinal fluid, a large informal healthcare sector and limited access to testing in the formal sector. JEV focused surveillance activities have detected the virus in some areas, in particular the North (Rangpur) and West (Rajshahi) but the burden elsewhere is unknown. This gap means we cannot develop a data informed strategy that targets specific areas for prioritisation.

To understand the underlying burden of JEV around Bangladesh we can use nationally representative serostudies^10^. By testing for JEV-specific IgG antibodies, we can identify the proportion of individuals that have ever been infected by JEV in their lifetime. These efforts have been boosted by the use of novel assays that limit cross-reactivity with other flaviviruses, such as dengue virus.^11^ The results of this testing can inform mathematical models that quantify the underlying burden around the country and the potential impact of different vaccination strategies.

## Methods

### National cross-sectional seroprevalence study

We conducted a nationally-representative community study in 70 communities across Bangladesh between October 2015 and January 2016 (N=2,938 individuals). The details of the study have been published elsewhere^10,12^. Briefly, 70 communities were randomly selected from a total of 97,162 listed in the national census. In each community, field teams approached at least 10 randomly selected households for participation. The field teams applied a structured questionnaire covering household as well as individual demographic data. Additionally, they collected approximately 5 mL of blood if above 3 years of age and 3mL if 3 years old or younger. The samples were tested for antibodies against IgG JEV using a bead-based multiplex serological assay. This assay uses the domain III of the flaviviruses E protein, which limits cross-reactivity across flaviviruses^11^. We considered individuals to be seropositive (i.e., had been infected with JEV) if they had a relative fluorescence intensity (RFI) of 3 or greater (mean RFI over a control)^6^.

### Mosquito trapping and speciation

In each community, eight of the participating households were randomly selected for participation for mosquito trapping. Participants who agreed were left a BG-Sentinel trap placed indoors and allowed to operate for 24 hours. The collected mosquitoes were transported to icddr,b laboratories where trained entomologists performed species identification through standard morphological keys.^13^ Mosquitoes were counted by species and sexes were documented for each community.

### Factors associated with serostatus

We used a spatially explicit logistic regression model to study individual-, household- and community-level factors associated with a JEV infection. We fit all models using a binomial generalised linear model with a logit link function in a hierarchical Bayesian framework of the individual serostatus data. We modelled a continuous spatial random effect as a zero-mean Gaussian process with a Matérn covariance function representing the residual spatial variation not explained by the factors included, computed with stochastic partial differential equations.^14,15^ We fit the model using an integrated nested Laplace approximation (INLA).^16^ Due to the possible collinearity between some covariates and the spatial effect, we accounted for spatial dependence constraining the effect of the spatial random intercept on the outcome.^6,17^ We tested whether individual factors were associated with risk, including age, sex, travel history, and education. We grouped individuals into seven age bins (0–9, 10–19, 20–29, 30–39, 40-49, 50-59 and 60+) and head of household education into four bins (no formal education, primary/middle school, high school, and higher education). Sex and travel history were self-reported upon enrollment into the survey. Covariates associated with the household being the number of household members, owning land outside the household. Community factors were living in an urban versus rural area, presence of pigs in the community, having seen nomadic pigs and detection of trapped mosquito species.

### Catalytic model

To estimate the underlying per capita risk of infection for susceptible individuals, or the force of infection^18^, across Bangladesh, we developed a spatially explicit serocatalytic model in a hierarchical Bayesian framework to fit the individual serostatus data. We considered two separate approaches. First, we assumed that the number of JEV seropositive individuals followed a zero-inflated binomial distribution and used log(age) as an offset and a cloglog link function (Model 1). This allowed us to separately estimate the probability (θ) that JEV circulates within a location (as one minus the probability of zero-inflation) - and, separately, the force of infection in the locations it circulates ^16,19,20^ This approach assumes that the force of infection in all places where JEV circulates is the same, since the model focuses on quantifying the probability of circulation within each location (θ). As an alternative, we implemented a second approach that assumed that serostatus followed a spatially varying binomial distribution and used log(age) as an offset and a cloglog link function (Model 2). This approach assumes that JEV can circulate everywhere but the magnitude differs. The key issue with this second approach is that a non-zero force of infection is estimated in all locations, which can lead to a substantial number of infections in large population centers, such as Dhaka, even if there is a very low force of infection estimated in that location. While neither model is perfect, by presenting the results of both analyses, we present a range of possible patterns of infection across the country. In both models, we assumed that JEV was first reported in 1977, consistent with that found elsewhere^6^. In both models, we modelled the spatial random effect using a Matern covariance function, as implemented by INLA.^16^ Probabilities and transmission parameters were estimated through the mean and 95% credible interval of the posterior distribution.

### Japanese encephalitis burden and health outcomes in Bangladesh

To estimate the annual JE disease burden, we estimated the number of infections, number of hospitalised cases, disability adjusted life years (DALYs) and deaths. To calculate the weighted sum of the number of infections at each age at each location we used the following expression derived from the serocatalytic model^21^:

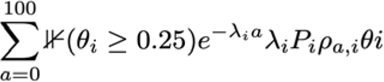

where:

θ_i_ represents one minus the probability of zero inflation, meaning the probability that JEV is present in cell *i*. This assumes that all locations with a low probability (<0.25) that JEV circulates (i.e., having non-zero seropositivity) are set to zero. This is to avoid large population centres (such as Dhaka) having a disproportionately large proportion of the national burden from JEV. In the alternative form of the model, θi is set to 1 for all locations.

λ_i_ represents the force of infection at cell *i* (this will be identical for all locations in the first model but varying across locations in the second model)

P_i_ represents the size of the population residing within cell *i*.

ρ_a_ represents the proportion of the population within cell *i* that are of age *a*., where individuals that were older than 38 were all given the value of 38. This is to reflect that the first report of JEV was in 1977 (38 years prior to the study) and therefore all individuals older than 38 would have experienced the same cumulative force of infection.

Using the number of annual infections, we estimated the number of people with disease using published estimates of the asymptomatic/symptomatic ratio, the proportions of severity levels (mild, moderate and severe) and the infection-fatality ratio (IFR) (Table 2).^1,2,8^ We also calculated the disability adjusted life years from JEV (DALY). DALYs indicate the health loss due to both nonfatal and fatal disease burden, calculated as the sum of years of life lost (YLLs) due to premature mortality and years lived with disability (YLDs). The YLL is based on remaining life expectancy when compared with a reference life table at age of death, and the YLD is calculated by multiplying the prevalence of a disease or injury and its main disabling outcomes by its weighted level of severity. As such, one DALY represents one year of healthy life lost. For the disability weights, we used the infectious disease for acute episodes and the motor and cognitive impairments at mild, moderate, and severe levels for long-term sequelae, from the Global Burden of Disease study 2013.^22^ For asymptomatic JE cases, we used no disability weight, since no burden is expected from that disease state.

### Japanese encephalitis vaccination scenarios

Since humans are incidental hosts for JEV, human infection happens through mosquito bites arising from zoonotic reservoirs.^2^ This means human vaccination has no impact on the underlying force of infection in a community and only those vaccinated are protected. To critically assess the impact of vaccination, we used the expected number of individuals by age and location that had each infection outcome (disease, severe disease and death) by year.

We developed a set of four different vaccination scenarios based on WHO recommendations^23^ and in consultation with immunization experts in Bangladesh, taking in consideration national epidemiological parameters such as geography, age groups and service delivery (vaccine coverage) (Table 3). All scenarios include a one-time multi-age cohort (MAC) campaign, in which we varied the geographical area, age target and vaccine coverage. All scenarios include the subsequent introduction of the JEV vaccine into a routine immunisation program at nine months of age, consistent with the target population of the measles-mumps-rubella (MMR) vaccine. In our simulations, we used a vaccine efficacy of 86.3%.^24^ Vaccine efficacy was considered to confer protection against disease and prevent vaccinated individuals who become infected from progressing to disease, and thus averting hospitalisation, long-term sequelae (DALYs) and death.

### Sensitivity analyses

To assess the robustness of our estimates, we conducted sensitivity analyses in which we used a spatially varying force of infection model and modified our assumption about the probability of severe disease (changing it from 1/1000 to 1/250). We also ran four additional vaccine scenarios where we changed the vaccine coverage (range 20-90), geographical (top 20-30 districts/zilas) and age groups targeted (1-90 years) for the initial MAC immunisation campaign. All analyses were conducted in R (version 4.3.2).

### Cost-effectiveness analysis

We conducted a model based cost-effectiveness analysis for JEV vaccination strategies in Bangladesh. The analysis adopted a public sector healthcare perspective and considered direct programmatic costs and health outcomes over a five-year time horizon, consistent with funding and implementation cycles. We presented the cost per DALY averted for each vaccine strategy and used the United States of America Dollar (US$) for currency costs. Programme costs were estimated per dose and included $0.45 per vaccine dose procurement and an additional $0.2 for delivery, monitoring and logistics. Health system costs per JE case were derived from a recent study in Bangladesh where acute JE episode (US$ 929), initial sequelae (US$ 75/month) and long-term sequence (US$ 47/month) were costed separately. We assumed that discounting begins at the age of vaccination, as protection from JEV vaccination is provided at the individual level. As the reference for a potential Willingness-To-Pay (WTP) threshold in Bangladesh, we used the GDP per capita and its multiples for 2023 (US$ 2,551)^25^. In the base case analysis, we applied a 0% discount rate for health effects and a 3% discount rate for costs. In sensitivity analysis, we ran two scenarios, one with an increase of ten times the force of infection and a second with an asymptomatic to symptomatic ratio of 1/250 (four times the base case) to assess the impact of higher burden scenarios.

### Ethical considerations

This study was approved by the icddr,b ethical review board (PR-14058). All adult participants provided written, informed consent after receiving detailed explanation of the study and procedures. Parents/guardians of all child participants were asked to provide written, informed consent on their behalf.

## Results

We assessed the presence of antibodies against JEV in 2,938 individuals with a mean age of 26 years ranging from 1-90 years of age (52% female) from 70 communities. The participants resided in all 8 divisions (administrative level 1), 62/64 zilas (administrative level 2), and 34 upazilas (administrative level 3) of the country’s administrative levels (Figure 1A). The overall JEV seroprevalence was 3.4% (95%CI 2.8-4.1) and was spatially heterogeneous, ranging from 0-28%, with the highest seroprevalence observed in a community in the north Mymensingh division near the border with India. In 23 (33%) communities, no individuals tested positive. Seroprevalence increased with age until age 38 years, consistent with the endemic transmission of JEV since 1977 (Figure 1B) and no differences by sex (p value: 0.25).

**Figure 1.**
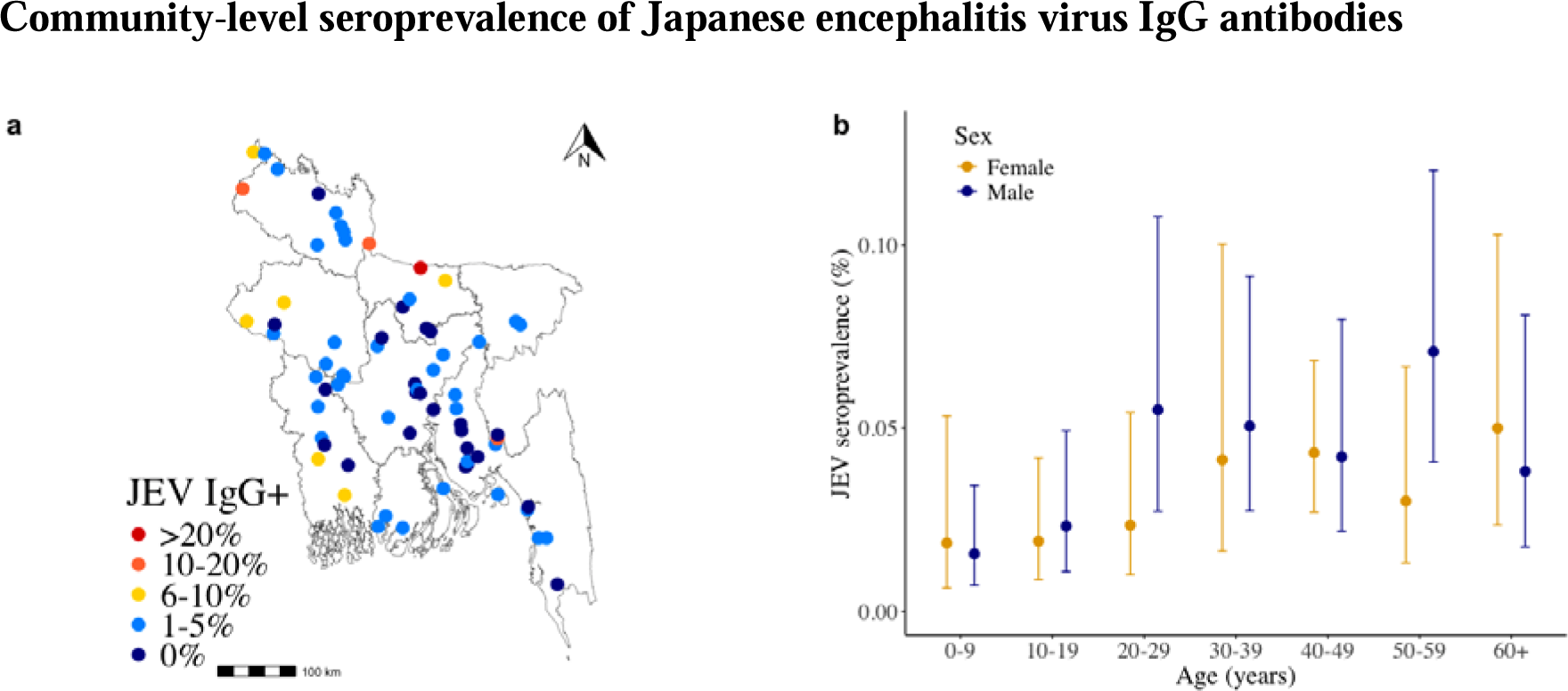
Japanese encephalitis seropositivity in sampled population. **a**, Location of the 70 sampled communities and JEV IgG seroprevalence at each community. **b**, Mean observed JEV IgG seropositivity by age group and sex (male - green, female - blue) and 95% confidence intervals.

We considered potential risk factors associated with being seropositive, and found that living in urban areas [odds ratio (OR) 0.53 (95% credible interval (CI): 0.29-0.94)] and population density were negatively associated with JEV seropositivity (adjusted (aOR) 0.81, 95%CrI: 0.70-0.94) (Table 1). Having contact with pigs (reported by 5.7% of individuals) was associated with being JEV seropositive (OR 2.09, 95%CrI: 1.00-4.30), as was reporting nomadic pigs passing through the community (OR 2.06, 95%CI: 1.25-3.38).

**Table 1.** Individual-, household-and community-level of participants stratified by serostatus to JEV.

|  | Overall, N=2938 | Seronegative,<br>n=2838 | Seropositive,<br>n=100 | Univariable<br>model,<br>unadjusted odds<br>ratio (95% CrI) |
| --- | --- | --- | --- | --- |
| **Individual-level risk factors** |  |  |  |  |
| Age |  |  |  |  |
| 0-9 | 352 (12.0) | 346 (12.2) | 6 (6.0) |  |
| 10-19 | 753 (25.6) | 737 (26.0) | 16 (16.0) |  |
| 20-29 | 498 (17.0) | 480 (16.9) | 18 (18.0) |  |
| 30-39 | 444 (15.1) | 424 (14.9) | 20 (20.0) |  |
| 40-49 | 374 (12.7) | 358 (12.6) | 16 (16.0)" |  |
| 50-59 | 260 (8.8) | 247 (8.7) | 13 (13.0) |  |
| 60+ | 257 (8.7) | 246 (8.7) | 11 (11.0) |  |
| Female | 1406 (47.9) | 1352 (47.6) | 54 (54.0) | 0.77 (0.51-1.15) |
| Travel last 6 months | 1526 (51.9) | 1468 (51.7) | 58 (58.0) | 1.29 (0.86-1.95) |
| Travel last 30 days | 977 (33.3) | 940 (33.1) | 37 (37.0) | 1.23 (0.81-1.87) |
| Travel last 7 days | 452 (15.4) | 434 (15.3) | 18 (18.0) | 1.24 (0.73-2.11) |
| **Household-level risk factors** |  |  |  |  |
| Number of household members | 6.22 (2.35) | 6.23 (2.35) | 6.00 (2.45) | 0.95 (0.86-1.04) |
| Education head of household |  |  |  |  |
| High school | 393 (13.4) | 373 (13.1) | 20 (20.0) | REF |
| Higher education | 474 (16.1) | 461 (16.2) | 13 (13.0) | 1 (0.98-1.02) |
| No formal education or illiterate | 908 (30.9) | 885 (31.2) | 23 (23.0) | 1 (0.98-1.02) |
| Primary/middle school | 1163 (39.6) | 1119 (39.4) | 44 (44.0) | 1 (0.98-1.02) |
| **Community-level risk factors** |  |  |  |  |
| Population density (log/km2) | 10.73 (1.19) | 10.74 (1.19) | 10.22 (0.93) | 0.62 (0.47-0.82) |
| Living urban area | 753 (25.6) | 739 (26.0)" | 14 (14.0) | 0.52 (0.29-0.94) |
| Any pigs in community | 1785 (60.8) | 1708 (60.2) | 77 (77.0) | 2.06 (1.25-3.38) |
| Mosquito species |  |  |  |  |
| *Cx. quinquefasciatus* | 2680 (91.2) | 2603 (91.7) | 77 (77.0) | 0.32 (0.19-0.54) |
| *Cx. tritaeniorhynchus* | 1125 (38.3) | 1081 (38.1) | 44 (44.0) | 1.11 (0.71-1.75) |
| *Cx. vishnui* | 752 (25.6) | 732 (25.8) | 20 (20.0) | 0.65 (0.38-1.12) |
| *Cx. pseudovishnui* | 1142 (38.9) | 1099 (38.7) | 43 (43.0) | 1.08 (0.69-1.67) |
| *Cx. fuscocephala* | 124 (4.2) | 119 (4.2) | 5 (5.0) | 1.38 (0.54-3.54) |
| *Cx. gelidus* | 627 (21.3) | 604 (21.3) | 23 (23.0) | 1.12 (0.67-1.87) |
| *Cx. bitaeniorhynchus* | 376 (12.8) | 359 (12.6) | 17 (17.0) | 1.19 (0.64-2.19) |
| *Ar. subalvatus* | 1736 (59.1) | 1675 (59.0) | 61 (61.0) | 0.90 (0.57-1.42) |
| *Ae. albopictus* | 1231 (41.9) | 1192 (42.0) | 39 (39.0) | 0.85 (0.55-1.33) |

\*n (%); mean (standard deviation).

**Table 2.** Summary of model parameters.

| Parameters | Estimate | Range | Reference |
| --- | --- | --- | --- |
| Asymptomatic to symptomatic severe cases | 1000:1 | 1000:1- 250:1 | 6 |
| Probability of sequelae | Uniform distribution | 0.3-0.5 | 3 |
| Long term sequelae severity |  |  | 30 |
| Mild | 0.15 | - |  |
| Moderate | 0.36 | - |  |
| Severe | 0.49 | - |  |
| Case fatality ratio | 0.2 | 0.2-0.3 | 34 |
| Duration of acute JE episode (days) | 20 | - | 35 |
| Disability weight for acute disease (per episode) |  |  | 22 |
| Severe | 0.133 | 0.148-0.308 |  |
| Disability weight for long term sequelae (annual) |  |  | 22 |
| Mild | 0.031 | 0.018-0.050 |  |
| Moderate | 0.203 | 0.134-0.290 |  |
| Severe | 0.542 | 0.374-0.702 |  |
| Vaccine efficacy | 0.95 | 0.67-0.99 | 36–38 |

**Table 3.** Vaccination scenarios for Japanese encephalitis vaccine introduction (campaign and routine immunization) in Bangladesh and correspondent geographical area, age target and vaccine coverage parameters.

| Scenarios | Geographical area | Age target | Vaccine coverage (%) |
| --- | --- | --- | --- |
| **A.** |  |  |  |
| Campaign | National | 1 – 15 years | 60 |
| Routine | National | 9 months | 90 |
| **B.** |  |  |  |
| Campaign | Top 10 zilas/districts | 1 – 15 years | 60 |
| Routine | Top 10 zilas/districts | 9 months | 90 |
| **C.** |  |  |  |
| Campaign | Top 10 zilas/districts | 1 – 90 years | 60 |
| Routine | Top 10 zilas/districts | 9 months | 90 |
| **D.** |  |  |  |
| Campaign | Top 3 divisions | 1 – 90 years | 60 |
| Routine | Top 3 divisions | 9 months | 90 |

Legend: Light-red fill shows changes related to base case scenario A.

We identified nine mosquito species as competent JEV vectors. There was substantial spatial heterogeneity wherein the different mosquito species were found (Table 1). The JEV vector species more commonly found in Bangladesh was *Culex* (*Cx.*) *quinquefasciatus* (present in 64 of 70 communities). In communities with JEV seropositive individuals *Cx. quinquefasciatus* (49/54, 70%), *Armigeres subalbatus* (31/54, 44%), and *Aedes albopictus* (25/54, 36%) were the most common mosquitoes captured. No species showed a positive association with JEV seropositivity in the sampled communities.

To estimate the national distribution in risk, we fitted a spatially-informed serocatalytic model to the serostatus data. We found a highly heterogeneous distribution of infection risk, with transmission concentrated in the border regions of the country, with little to no transmission in the centre of the country (Figure 2a). We estimated an overall average force of infection of 0.005 (95%CI:0.003-0.009), meaning 0.5% of the population gets infected with JEV each year. We estimated that there were an average of 157,000 annual JEV infections nationally (95% credible interval (CrI): 88,000–261,000) (Figure 2b). Every year these infections lead to an estimated 157 (95%CrI 89–253) hospitalised cases (Figure 2c), 31 (95%CrI 18-52) deaths (Figure 2d), and 112 (95%CrI 38–303) disability-adjusted life years lost (DALYs). We estimated that the mean age of cases was 30 years. In sensitivity analysis, the number of infections varied with assumptions about the probability of transmission, force of infection model and the probability of severe disease (Table 4). With the mean number of annual cases ranging from 156-628 across the different assumptions considered, and the mean number of deaths ranging from 31-114.

**Figure 2.**
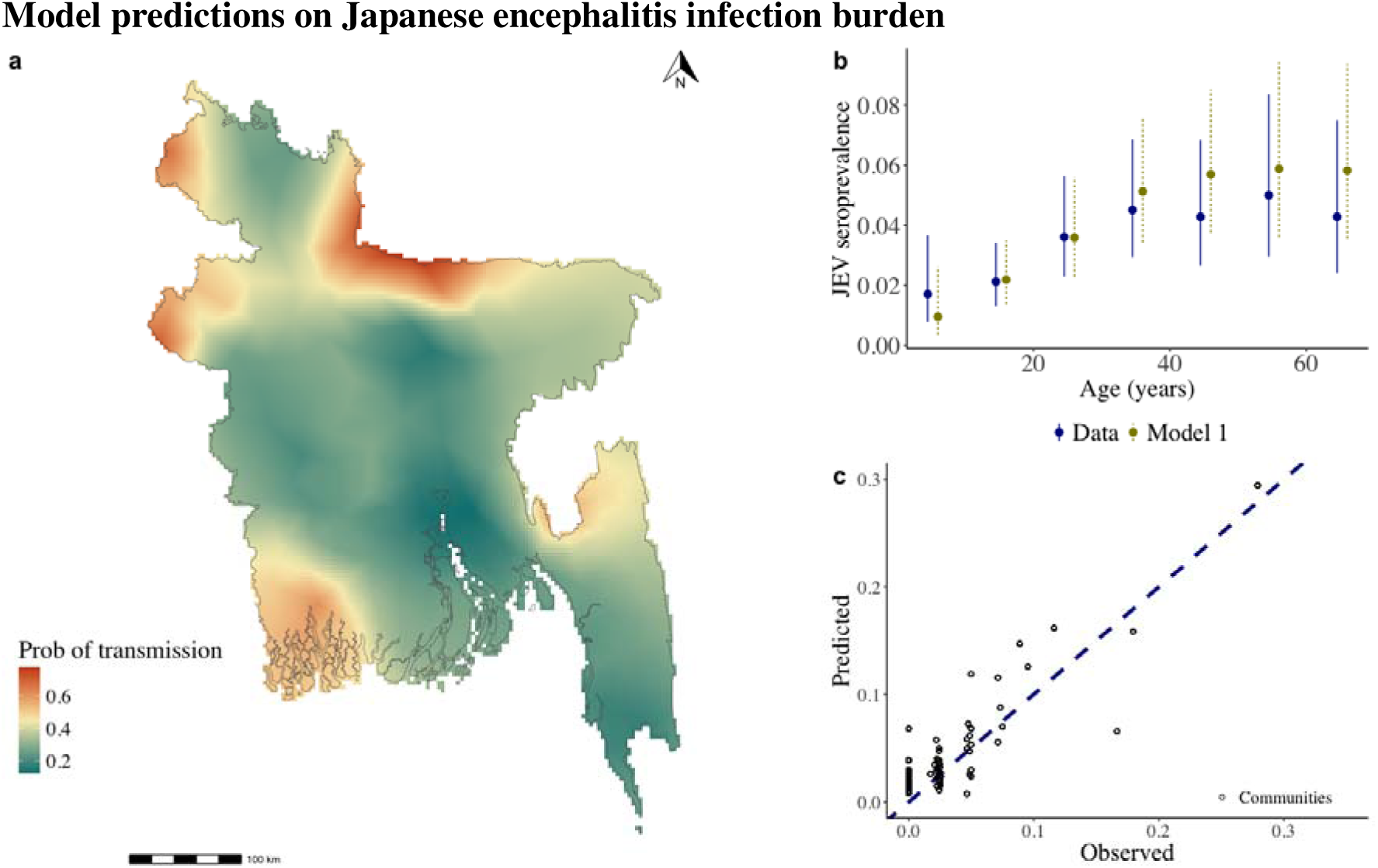
Japanese encephalitis model predictions. **a,** Predicted probability of JEV transmission (θ) in Bangladesh. **b,** Data and model JEV seroprevalence for each individual sampled by age group using a zero-inflated binomial serocatalytic spatial model. **c**, Observed and predicted JEV seroprevalence in each sampled communities (back open circles) using a zero-inflated binomial serocatalytic spatial model. Blue dashed line represents *y=x*.

**Table 4.** Summary of sensitivity analysis on health outcomes estimated using different models and/or parameters.

|  | **Model 1**<br>**Full penalised transmission**<br>**with $\theta$** | **Model 2**<br>**Spatially varying force of**<br>**infection ($\lambda$)** |
| --- | --- | --- |
| **Force of infection ($\lambda$)** | 0.005 (95% CI: 0.003-0.009) | 0.001 (95% CI: 0.0005-0.003) |
| **Infections** | 157,000 (95% CI: 88,000-253,000) | 156,000 (95% CI: 42,000-559,000) |
| **Cases (hospitalisations)**<br>**1:1000** | 157 (95% CI: 89-253) | 156 (95% CI: 42-559) |
| **Cases (hospitalisations)**<br>**1:250** | 628 (95% CI: 355-1,013) | 624 (95% CI: 168-2,236) |
| **Disability-adjusted life years**<br>**(DALYs)** | 113 (95% CI: 39-281) | 121 (95% CI: 11-1,626) |
| **Deaths (case fatality ratio 20%)** | 31 (95% CI: 18-52) | 42 (95% CI: 11-146) |
| **Deaths (case fatality ratio 30%)** | 47 (95% CI: 26-78) | 47 (95% CI: 12-166) |
| **Mean age of infection** | 30 years | 31 years |

Among the eight divisions of Bangladesh, the annual case incidence was highest in Rangpur (0.17 cases per 100,000 population, 95%CI 0.24-0.71), followed by Mymensingh and Sylhet (0.16 cases per 100,000 population, 95% CI 0.23-0.67) (Supplementary Figure 2). At the district level, the annual JE case incidence was highest in the Sherpur district - Mymensingh division, with 0.26 (95%CI 0.15-0.43) cases per 100,000 population, followed by Chapai Nawabganj (0.24 cases per 100,000, 95%CI 0.14-0.40) (Supplementary Figure 2). We also mapped the 10, 20 and 30 districts, as well as the three divisions with the highest JE case incidence to support vaccine strategy planning (Supplementary Figure 3).

We next modelled the impact of different vaccination strategies, starting with a one-time multi-age cohort (MAC) campaign, followed by the introduction of the JEV vaccine into the Expanded Programme on Immunization (EPI) targeting nine-month-old children (Figure 3). The number of cases averted over a five-year horizon (one-year campaign + four-year EPI) ranged from 26 to 348 over five years across the four scenarios considered and required between 5 and 35 million doses. We estimated that a national campaign (scenario A) targeting cohorts between 1 to 15 years of age with a 60% population covered would require 35 million doses over 5 years and prevent 167 cases, 33 deaths and 100 DALYs (0.5 cases averted per 100,000 doses used). Focusing just on the 10 most affected districts (scenario B) with the same initial target age group would require 5 million doses over 5 years and prevent 49 cases, 10 deaths and 29 DALYs (1 case averted per 100,000 doses used). Initially expanding the campaign to individuals of all ages (scenario C) in the top 10 districts would increase the impact to 135 cases, 27 deaths and 80 DALYs (0.9 cases averted per 100,000 doses used). Focusing on three highest incidence divisions, Rangpur, Mymensingh and Sylhet (scenario D) and individuals of 1-15 ages and 60% coverage would require 26 million doses and result in 186 cases, 37 deaths and 111 DALYs averted (0.7 case averted per 100,000 doses used). We also tested four alternative scenarios, varying assumptions and model (binomial varying FOI), the targeted districts, age groups and vaccine coverage (Supplementary Tables 1-3) and found that 3-51 million doses would be needed to avert avert 26-348 cases, 5-70 deaths and 15-207 DALYs (maximum 1 case averted per 100,000 doses). Finally, a currently proposed strategy focusing on Rajshahi, Rangpur and Chittagong divisions would require 46 million doses and result in 240 cases, 48 deaths and 136 DALYs averted (0.5 case averted per 100,000 doses used).

**Figure 3.**
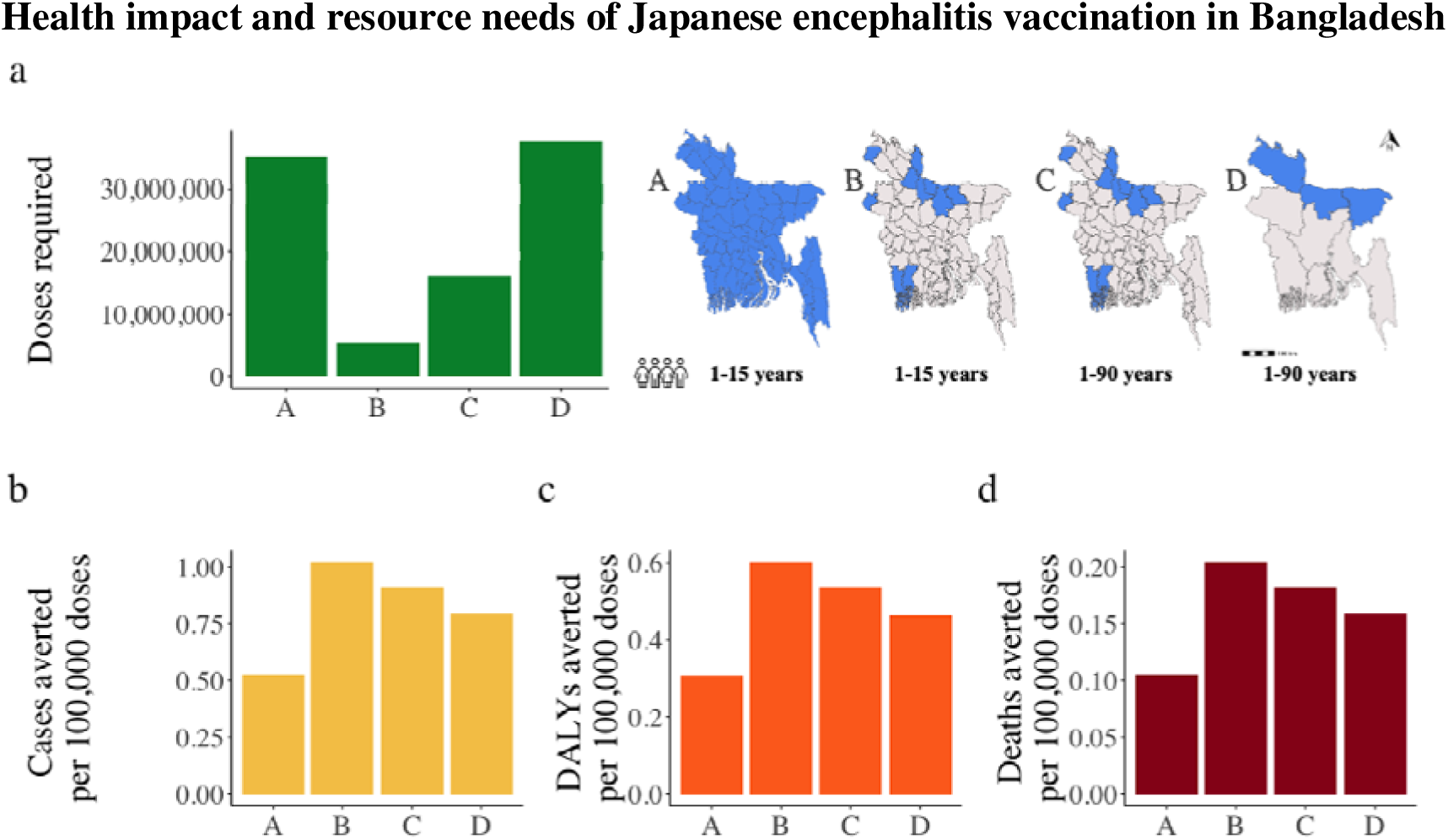
Projected 5-year health impact of Japanese encephalitis vaccination across four national and subnational scenarios. Impact of vaccination on the health burden of Japanese encephalitis for eight alternative vaccination scenarios, varying geographical area, age group and vaccine coverage of a one-time MAC campaign. Top row has cumulative doses required for each vaccination scenario (A-D) in a 5-year period (green bar plot). Bottom row has the cumulative number of cases (yellow bar plot), DALYs (orange bar plot) and deaths (red bar plot) averted per 100,000 doses for each vaccination scenario in a 5-year period.

Across all vaccination strategies, the cost per DALY averted ranged from approximately USD 108,000 to 210,000 over a five-year period (Supplementary Table 4). None of these scenarios met the cost-effectiveness thresholds of one- or three-times Bangladesh’s GDP per capita (USD 2,700 and USD 8,100, respectively). Estimated net savings were negative in all cases, with additional program costs ranging from USD 2 million to 33 million over a five-year period. The average cost per symptomatic case averted was estimated at USD 14, reflecting the weighted cost of acute care, sequelae, and mortality. The cost-benefit ratio across all strategies remained below 1 (range: 0.00013–0.00044), indicating that the program costs more than the direct illness costs saved. Incremental cost-effectiveness ratios (ICERs) against the base case scenario similarly exceeded acceptable benchmarks. Sensitivity analyses showed that increasing the asymptomatic-to-symptomatic ratio to 1:250 improved cost-effectiveness, with cost per DALY averted decreasing to a quarter of the initial calculations to USD 24,000–47,000; however, no scenarios reached cost-effectiveness under standard WTP thresholds (Supplementary Table 5). When we increased the FOI to 0.05 (10 times more than estimated by our data), the cost per DALY averted decreased, ranging from USD 12,000 to 39,000 across strategies but remained under the WTP thresholds (Supplementary Table 5).

## Discussion

In this study we have estimated the national burden from JEV in Bangladesh using a representative serological study and projected the impact of alternative Japanese encephalitis vaccination scenarios. We found that around 5 million individuals (around 3% of the population) have been infected with JEV in their lifetime, most subclinically, with around 157,000 infections and 41 deaths per year. The risk of infection varied greatly across the country, with the highest prevalence found in the border regions. Our estimates can form the basis of planning the distribution of vaccines in the country.

The underlying risk of JEV infection in Bangladesh is lower than other countries in the region. This is likely due to Bangladesh being predominantly a Muslim country, with only a few communities raising pigs, typically as single backyard pigs or through nomadic pig herds.^26^ We found that having pigs in the community was one of the significant risk factors associated with seropositivity. Many areas on the Indian side of the border where we saw higher levels of JEV risk have large pig populations, for example the Meghalaya state, belonging to the North East Hill region of India which combined has 46% of India’s total pig population.^27^ We also found that low population density was positively associated with a past JEV infection, which was expected since rural areas with rice fields and flooded areas are suitable ecological niches for JEV transmission.^28^ We found several JEV competent mosquito vector species throughout the country, suggesting that the potential for transmission exists over a much wider footprint than currently observed.

Despite the low number of cases nationally, a geographically targeted vaccine campaign could have an important impact on the burden of Japanese encephalitis. Bangladesh has a strong track record of introducing new vaccines. Bangladesh has recently successfully deployed two vaccines to age groups not previously covered by the EPI: the tetanus-diphtheria vaccine for adults and the human papillomavirus virus vaccine for girls and female adolescents^29^. As children between 9 and 15 months of age are eligible for the MMR vaccine, the introduction of the JE vaccine alongside the MMR vaccine schedule, at 9 months of age, would maximise vaccine uptake and coverage. However, only targeting the vaccine in the youngest age groups through the EPI would take several decades to see the full public health benefit of this approach given the mean age of infection. We estimated that the mean age of cases is 31 years. Initial supplementary immunisation activities that target a MAC will be important to maximise the impact of the vaccine. Ultimately, the optimal geographical unit and age group to roll out vaccines will depend on logistical and budgetary constraints. Most vaccines are managed at the national or at the division level and it may not be feasible to roll out vaccines at a finer spatial scale.

Our results differ from previous estimates obtained from hospital catchment areas in Rajshahi, Khulna and Chittagong, where JE incidence was estimated to be 2.7, 1.4 and 0.6 per 100,000 population, respectively.^5^ Our results are also different from a recent cost-effectiveness analysis, where JE case incidence was assumed to be 2.5 per 100,000 population for the whole country.^7,30^ Our JE case incidence estimates for Rajshahi, Khulna and Chittagong are around ten times lower at 0.15, 0.15 and 0.10 per 100,000 population, respectively. The reasons for these discrepancies are unclear. Our infection estimates are based on population representative samples at a national level, and are therefore unbiased by healthcare seeking behaviours or access to testing. We note that the inferred mean age of cases (31 years) is very similar to that observed in the country (30 years)^31^, suggesting that we are accurately capturing the correct underlying force of infection. Countries such as Nepal had a much lower mean age of cases prior to the introduction of the vaccine (14 years), consistent with a higher force of infection. Our estimates of the number of cases are reliant on translating underlying levels of infection to numbers of severe cases and deaths, reliant on a previous study in Bangladesh that linked underlying infection from a seroprevalence study in west Bangladesh with levels of severe disease in the same area, which resulted in an estimate of around 1 severe case per 1,000 infections.^6^ If instead, the probability of severe disease was 1:250 infections, which has also been suggested, our estimates would increase to 1 case per 100,000. Using this higher probability of severe disease given disease increases slightly the potential impact of the vaccine (2-4 cases per 100,000 averted as compared to 1 case in the base case for scenario A).

In 2015, the same year as our seroprevalence study, the WHO Data Observatory reported 76 JE cases in Bangladesh, around 48% of the national JE case burden we estimated^32^. The distribution of risk burden is different to that reported through the surveillance activities in the country. This is likely due to the focused nature of these surveillance activities, where a few dedicated hospitals have the ability to do the necessary cerebrospinal fluid testing.^33^ However, these surveillance hospitals may not capture all cases, especially as they are all in urban environments, often far from the rural communities where JE cases occur. Our findings suggest that, in particular, surveillance for Japanese encephalitis should be expanded to communities in Sylhet, Mymensingh and Khulna which have some places with the highest burden but where few cases are detected.

Although JE vaccination can avert a substantial disease burden, no vaccination scenario considered was cost-effective when considering program costs under a WTP of one up to three GDP per capita, as per WHO-CHOICE standards. ICERs against the base case scenario similarly were very high, highlighting the challenges of achieving economic efficiency of JEV vaccination. Additionally, net savings from a vaccination program over 5 years were negative, when considering treatment of symptomatic cases under a public healthcare perspective. The cost-benefit ratios across all strategies remained well below 1 indicating that the direct medical cost savings from illness averted were substantially smaller than any vaccination program costs. The method of using WTP of a GDP product has several limitations and does not discriminate between good and bad value for money. It is a general consensus that under a public health perspective any preventive measure is better protecting potential healthy years than a treatment-palliative approach, since virtually most burden would be avoided. These results highlight that the economic justification for JEV vaccination, under the modeled assumptions, would rely on broader societal benefits beyond direct healthcare cost savings, such as equity impact on protecting vulnerable and minority populations, which may strengthen the economic case for JE vaccination in Bangladesh.

Our study is subject to limitations. While we used an assay that minimised cross reactivity with other flaviviruses, some cross-reactivity, especially with dengue virus, would lead to some false positives. This would mean that the true force of infection is lower than we report. Nevertheless, the effect is likely to be minimal - it has previously been shown that dengue at the time of this study was concentrated in urban hubs and therefore occupies a separate ecological niche than JEV^10^. We note that the serostudy is a few years old, and the epidemiology of JEV may have since changed. However, as humans are dead end hosts, this would require a big change in the underlying patterns of transmission in reservoirs or the size of the population at risk. We note the steady increase in seropositivity by age in individuals under 38 years consistent with stable transmission since it was first introduced in 1977. While we attempted multiple models to capture the true spatial heterogeneity in risk, our sampling approach was not powered to accurately capture spatial heterogeneity in risk across the country. This means our estimates of the force of infection at the district level should be taken with caution. Increased sampling of communities in the provinces with the highest risk will aid refine these estimates, and help identify specific districts that should be included in targeted vaccine campaigns. Only BG traps were available during the study period and the inclusion of new generation traps, such as light traps, could have given better understanding of the mosquito diversity in the study areas.

Although many countries are investing in their national immunisation programmes, new vaccines and higher target coverage rates are driving costs up. Comprehensive health impact analyses assist decision-makers with critical estimates on the outcomes averted through vaccine interventions in the context of competing public health priorities. Nationally representative seroprevalence studies, such as we present here, can underpin the underlying burden, including how it varies over space, allowing policy makers and funders to make evidence-based decisions on whether to invest in specific vaccines.

## Figures and tables

**Supplementary Figure 1.**
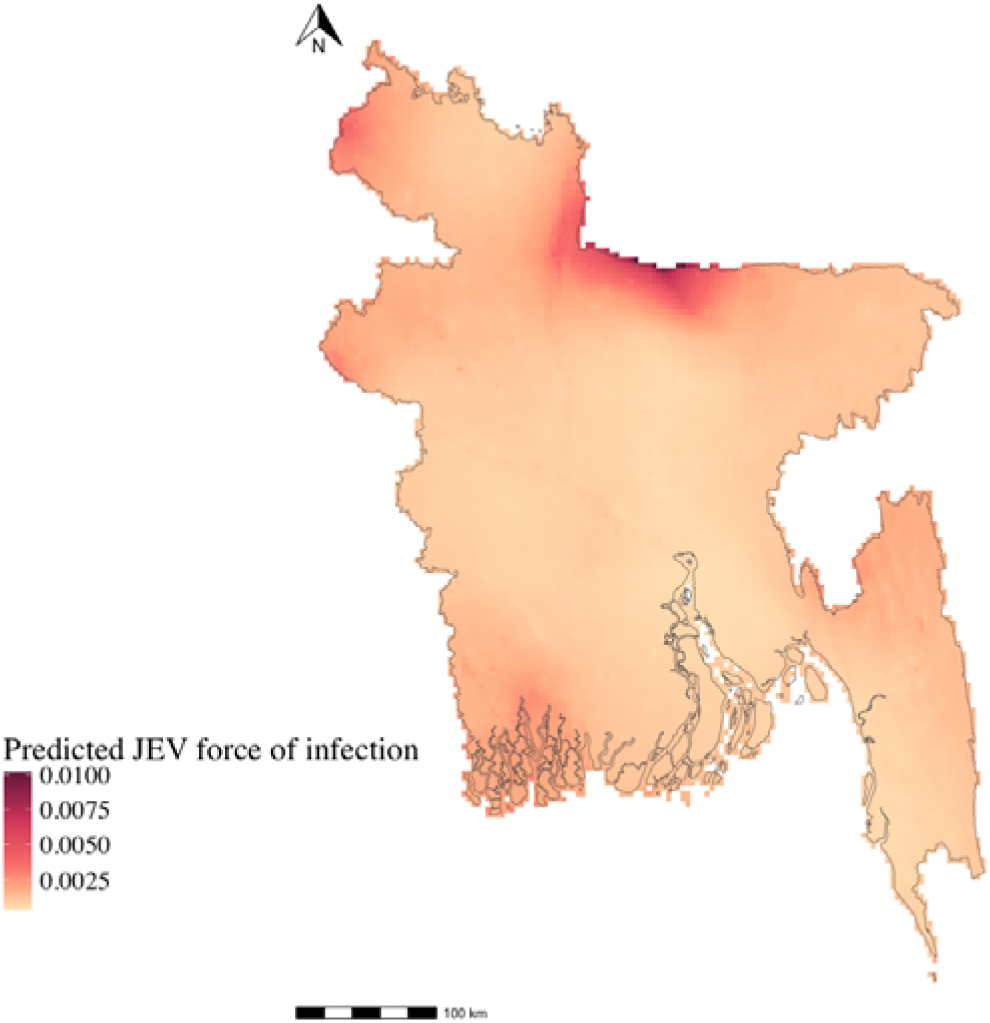
Predicted JEV force of infection using Model 2. Spatially explicit predicted JEV force of infection map using a Matérn covariance structure in a binomial model with a complementary log link function using the logarithm of age as an offset.

**Supplementary Figure 2.**
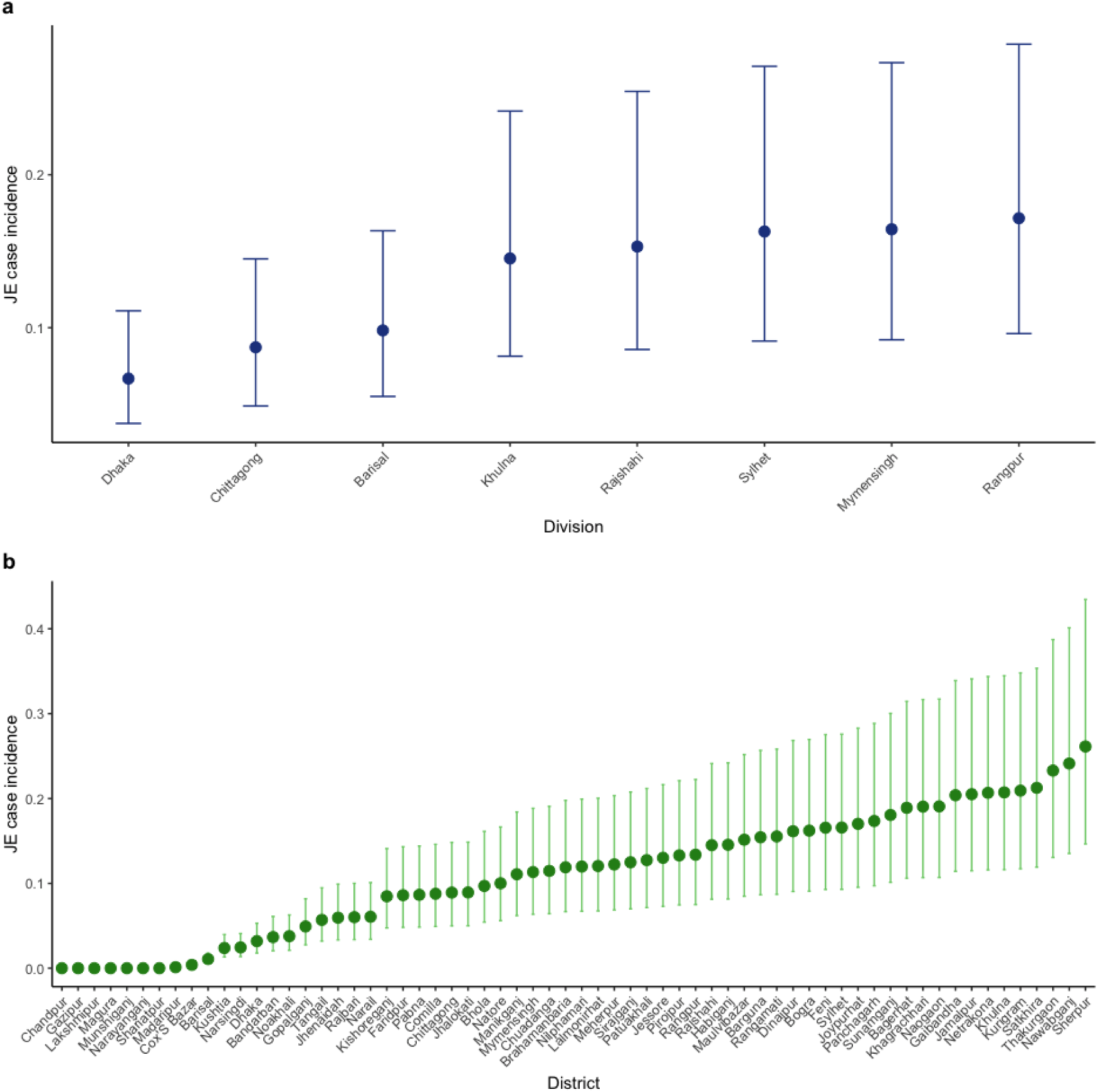
Japanese encephalitis case incidence per 100,000 population (ascendent) at different subnational administrative levels. **a**, JE case incidence at each eight subnational divisions (administrative level 1) of Bangladesh. **b**, JE case incidence at each 64 zilas/districts (administrative level 2) of Bangladesh.

**Supplementary Figure 3.**
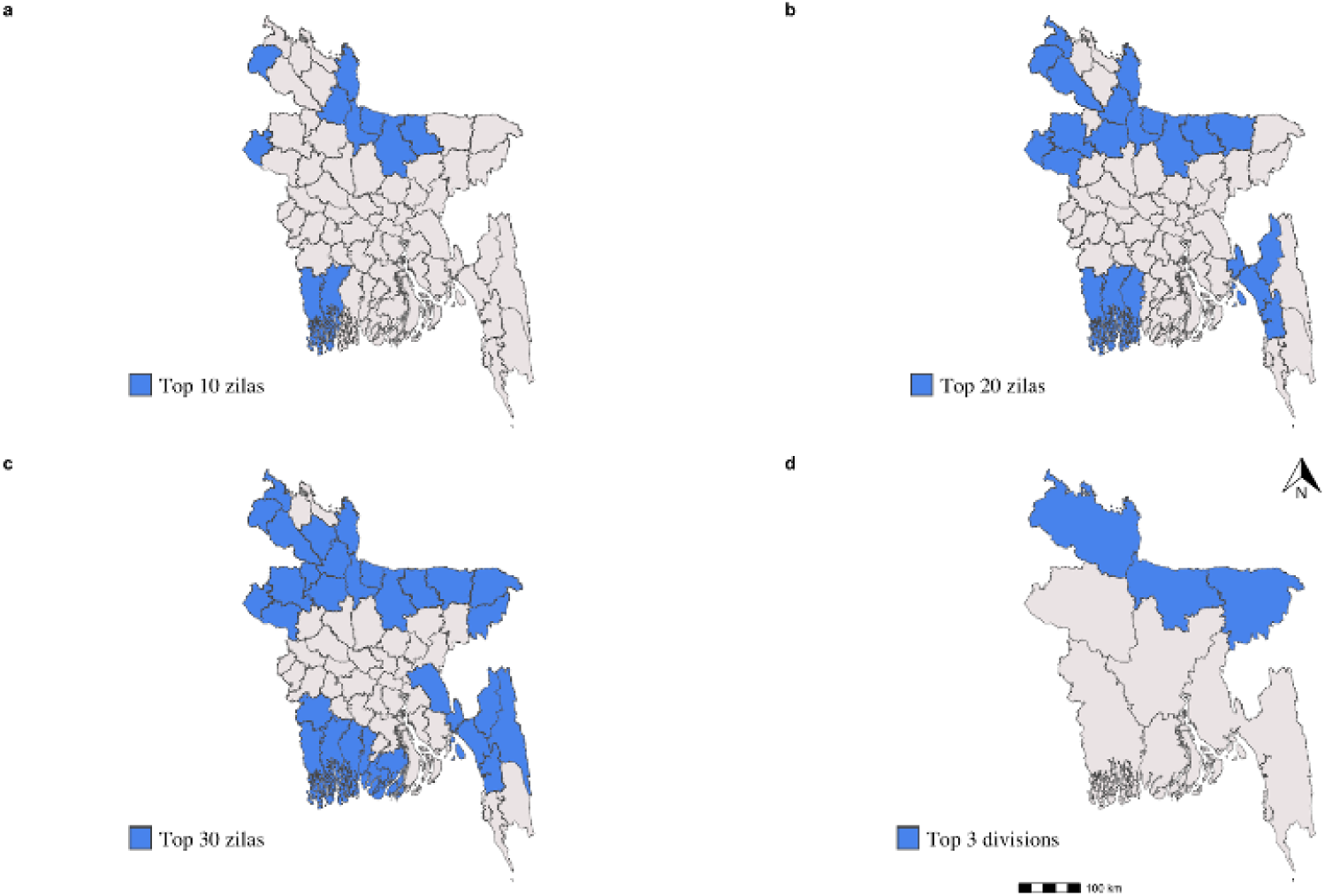
Geographical targets (zilas/districts and divisions) for proposed vaccination scenarios obtained from estimated JE case incidence. **a**, top 10 zilas/districts. **b**, top 20 zilas/districts. **c**, top 30 zilas/districts. **d**, top 3 divisions.

**Supplementary Table 1.** Additional vaccination scenarios for Japanese encephalitis vaccine introduction (campaign and routine immunization) in Bangladesh and correspondent geographical area, age target and vaccine coverage parameters.

| Scenarios | Geographical area | Age target | Vaccine coverage (%) |
| --- | --- | --- | --- |
| **E.** |  |  |  |
| Campaign | Top 10 zilas/districts | 1 – 15 years | 20 |
| Routine | Top 10 zilas/districts | 9 months | 90 |
| **F.** |  |  |  |
| Campaign | Top 20 zilas/districts | 1 – 15 years | 60 |
| Routine | Top 20 zilas/districts | 9 months | 90 |
| **G.** |  |  |  |
| Campaign | Top 20 zilas/districts | 1 – 90 years | 60 |
| Routine | Top 20 zilas/districts | 9 months | 90 |
| **H.** |  |  |  |
| Campaign | Top 30 zilas/districts | 1 – 90 years | 60 |
| Routine | Top 30 zilas/districts | 9 months | 90 |

Legend: Light-red fill shows changes related to base case scenario A.

**Supplementary Table 2.**
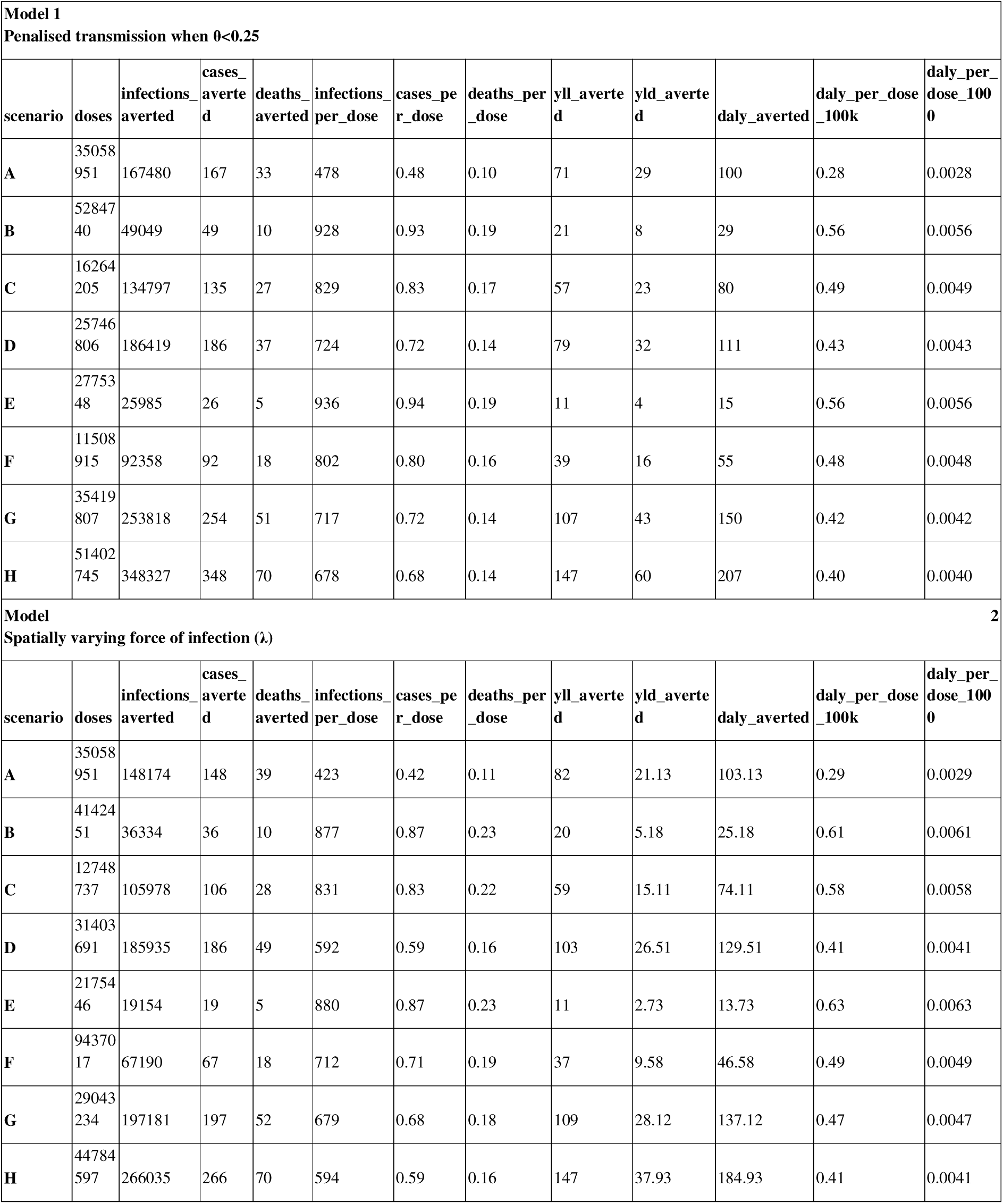
Impact of 8 vaccination scenarios, starting with a one-time multi-age cohort (MAC) campaign, followed by the introduction of the JEV vaccine in the Expanded Programme on Immunization (EPI) targeting one year old children (1+4 years horizon), by JE burden estimating model.

**Supplementary Table 3.** Model 1 and 2 burden estimates and comparison with published literature estimates.

|  | Force of infection | infections | Symptomatic/asymptomatic ratio | Severe cases | Case incidence | Mean age cases | Population<sup>39</sup> |
| --- | --- | --- | --- | --- | --- | --- | --- |
| **Chapai Nawabganj district paper** | 0.007<sup>6</sup> | 13,750<sup>6</sup> | 1/1000<sup>6</sup> | 12<sup>6</sup> | 0.70<sup>6</sup> | 31 | 1,886,583 |
|  | 0.007<sup>6</sup> | 13,750<sup>6</sup> | 1/250 | 55 | 2.90 | 31 | 1,886,583 |
| **Rajshahi catchment area:**<br>Rajshahi, Naogaon, Chapai Nawabganj districts | 0.79 | 181,000 | 1/1000 | 181 | 2.70<sup>5</sup> | 30 | 6,700,000 |
|  | 0.007 | 45,250 | 1/250 | 181 | 2.70<sup>5</sup> | 30 | 6,700,000 |
| **Model 1 - Rajshahi catchment area:**<br>Rajshahi, Naogaon, Chapai Nawabganj districts | 0.005 | 13,418 | 1/1000 | 13 | 0.18 | 30 | 7,688,915 |
|  | 0.005 | 13,418 | 1/250 | 54 | 0.70 | 30 | 7,688,915 |
| **Model 1 - Chapai Nawabganj district** | 0.005 | 4,445 | 1/1000 | 4 | 0.23 | 30 | 1,886,583 |
|  | 0.005 | 4,445 | 1/250 | 18 | 0.95 | 30 | 1,886,583 |
| **Model 2 - Rajshahi catchment area:**<br>Rajshahi, Naogaon, Chapai Nawabganj districts | 0.0016 | 10,513 | 1/1000 | 11 | 0.15 | 31 | 7,688,915 |
|  | 0.0016 | 10,513 | 1/250 | 42 | 0.55 | 31 | 7,688,915 |
| **Model 2 - Chapai Nawabganj district** | 0.002 | 3,473 | 1/1000 | 3 | 0.190 | 31 | 1,886,583 |
|  | 0.002 | 3,473 | 1/250 | 14 | 0.740 | 31 | 1,886,583 |

Legend: Cells in blue colour are values published in references <sup>5,6</sup>

**Supplementary Table 4.**
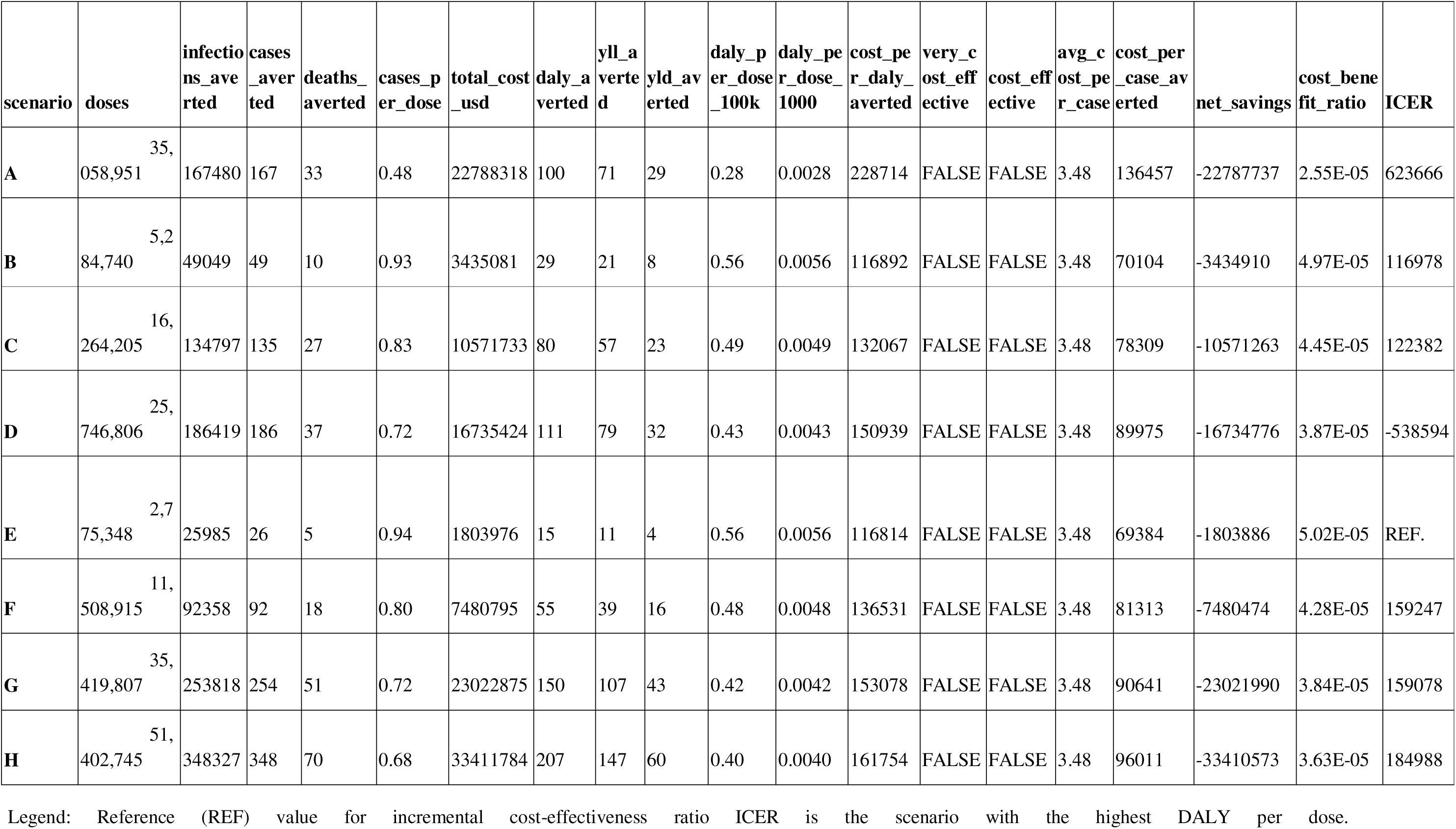
Cost-effectiveness analysis of JE vaccine scenarios.

**Supplementary Table 5.**
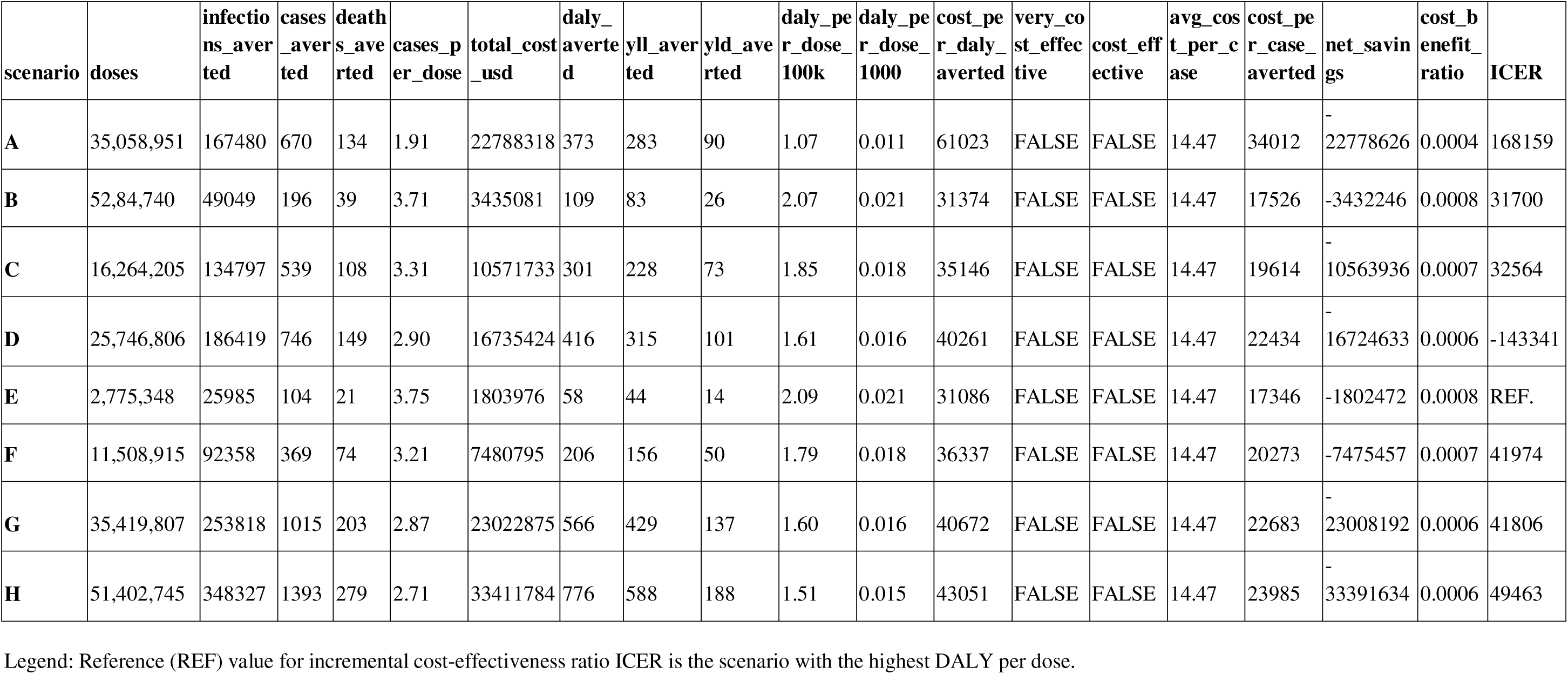
Sensitivity and cost-effectiveness analysis of JE vaccine scenarios under a 1/250 asymptomatic-to-symptomatic ratio.

**Supplementary Table 6.** Sensitivity and cost-effectiveness analysis of JE vaccine scenarios under a 0.05 force of infection.

| scenario | doses | Infections_aver_ted | Cases_aver_ted | Deaths_aver_ted | cases_per_dose | Total_Cost_usd | Daly_aver_ted | Yll_aver_ted | Yld_aver_ted | daly_per_dose_100k | daly_per_dose_1000 | cost_per_daly_aver_ted | very_cost_effective | Cost_effective | avg_cost_per_case | Cost_per_case_aver_ted | Net_savings | Cost_Benefit_ratio | ICER |
| --- | --- | --- | --- | --- | --- | --- | --- | --- | --- | --- | --- | --- | --- | --- | --- | --- | --- | --- | --- |
| **A** | 35,058,951 | 347789 | 348 | 87 | 0.99261384 | 22788318 | 909.903095 | 734 | 175.903095 | 2.60 | 0.02595352 | 25045 | FALS E | FALS E | 3.48 | 65484 | -22783430 | 0.00021449 | -150046.64 |
| **B** | 5,284,740 | 81457 | 81 | 20 | 1.53271495 | 3435081 | 213.198941 | 172 | 41.1989407 | 4.03 | 0.04034237 | 16112 | FALS E | FALS E | 3.48 | 42408 | -3433943 | 0.0003312 | 24354.6125 |
| **C** | 16,264,205 | 110752 | 111 | 28 | 0.68248033 | 10571733 | 290.015629 | 234 | 56.0156289 | 1.78 | 0.01783153 | 36452 | FALS E | FALS E | 3.48 | 95241 | -10570174 | 0.00014747 | 92904.9718 |
| **D** | 25,746,806 | 182326 | 182 | 46 | 0.7068838 | 16735424 | 477.215992 | 385 | 92.2159921 | 1.85 | 0.01853496 | 35069 | FALS E | FALS E | 3.48 | 91953 | -16732868 | 0.00015275 | 233203.663 |
| **E** | 2,775,348 | 55807 | 56 | 14 | 2.01776498 | 1803976 | 146.225804 | 118 | 28.2258036 | 5.27 | 0.05268738 | 12337 | FALS E | FALS E | 3.48 | 32214 | -1803190 | 0.00043601 | REF. |
| **F** | 11,508,915 | 167132 | 167 | 42 | 1.45104903 | 7480795 | 437.531242 | 353 | 84.5312418 | 3.80 | 0.03801672 | 17098 | FALS E | FALS E | 3.48 | 44795 | -7478449 | 0.00031355 | -20953.297 |
| **G** | 35,419,807 | 227239 | 227 | 57 | 0.64088435 | 23022875 | 594.931879 | 480 | 114.931879 | 1.68 | 0.01679659 | 38698 | FALS E | FALS E | 3.48 | 101422 | -23019686 | 0.00013849 | 53412.0822 |
| **H** | 51,402,745 | 320502 | 321 | 80 | 0.62448027 | 33411784 | 839.102004 | 677 | 162.102004 | 1.63 | 0.01632407 | 39819 | FALS E | FALS E | 3.48 | 104087 | -33407276 | 0.00013494 | 42547.8331 |

Legend: Reference (REF) value for incremental cost-effectiveness ratio ICER is the scenario with the highest DALY per dose.

## Acknowledgements

HS was supported by the European Research Council No. 101170844. S.C. acknowledges support from the European Commission under the EU4Health program 2021–2027, Grant Agreement-Project: 101102733—DURABLE, the Investissement d’Avenir program, the Laboratoire d’Excellence Integrative Biology of Emerging Infectious Diseases program (grant ANR-10-LABX-62-IBEID), the European Union’s Horizon 2020 research and innovation program under VEO grant agreement No. 874735, U01 NIH-PICREID (U01AI151758) and the INCEPTION project (PIA/ANR-16-CONV-0005). MPD is funded by Gates Cambridge Trust.

## Author contributions

Conceptualization: HS, EG, MPD

Methodology: MPD, GRS, MOD, EK, JV, SL, EG, HS

Investigation: AMN, KKP, MR, MSA, HMA, MZR, MEH, RCP, SL, EG, HS

Visualization: MPD

Supervision: HS, EG

Writing—original draft: MPD, HS

Writing—review & editing: All authors

## Data availability

The data and code to replicate main analyses are available at [will be provided upon acceptance]. To protect privacy, we grouped ages into age groups and showed the above sampling administrative level.

